# Five-Domain Accelerometer-Derived Behavioral Exposome and Incident Cancer Risk in UK Biobank

**DOI:** 10.64898/2026.04.07.26350369

**Authors:** Maung Ni Chan Chin (Chengqin Ni), Javier Alejandro Berrio

## Abstract

**Background:** Accelerometer-derived behavioral phenotype captures multidimensional aspects of human behavior extending well beyond physical activity, encompassing light exposure, step counts, physical activity patterns, sleep, and circadian rhythms. Whether these five domains constitute a unified behavioral architecture underlying cancer risk and whether circadian organization and light exposure confer incremental predictive value beyond movement volume alone remains to be comprehensively established.

**Methods:** We conducted an accelerometer-wide association study (AWAS) encompassing the complete accelerometer-derived behavioral exposome across five behavioral domains in UK Biobank participants with valid wrist accelerometry data. Incident solid cancers were designated as the primary endpoint, with prespecified site-specific solid cancers and hematological malignancy as secondary outcomes. Cox proportional hazards models with age as the timescale were used. The minimal covariate set served as the primary reporting tier, followed by sensitivity analyses additionally adjusting for adiposity/metabolic factors, independent activity patterns, shift work history, and accelerometry measurement quality. Nominal statistical significance was defined as two-sided P < 0.05

**Results:** Among 89,080 participants, 6,598 incident solid cancer events were observed over a median follow-up of 8.39 years. In the minimally adjusted model, the pan-solid-tumor association atlas was dominated by signals from activity volume, inactivity fragmentation, and circadian rhythm. Higher overall acceleration (HR per SD: 0.91, 95% CI: 0.89-0.94) and higher daily step counts (HR: 0.93, 95% CI: 0.90-0.95) were independently associated with reduced solid cancer risk, while inactivity fragmentation metrics were consistently linked to higher risk. Notably, circadian rhythms, most prominently cosinor mesor (Midline Estimating Statistic of Rhythm under cosinor model), emerged as leading inverse risk signals, underscoring the independent contribution of circadian behavioral architecture. Site-specific analyses revealed pronounced heterogeneity across tumor sites. Lung cancer exhibited a robust inverse activity-risk gradient, while breast cancer showed reproducible associations with MVPA. Most strikingly, nocturnal light exposure demonstrated a tumor-site-specific association confined to pancreatic cancer, a signal absent across all other sites examined. Associations for uterine cancer were predominantly inactivity-related and substantially attenuated following adjustment for adiposity and metabolic factors.

**Conclusions:** Across five accelerometer-derived behavioral domains, solid cancers as a whole were most consistently associated with a high-movement, low-fragmentation, and circadian-coherent behavioral profile. While site-specific heterogeneity exists, the broad cancer risk landscape is dominated by movement volume, inactivity fragmentation, and circadian rhythmicity. Light exposure, although more localized in its contribution, demonstrates a potentially novel and specific association with pancreatic cancer risk. These findings support a five-domain behavioral exposome framework for cancer epidemiology and, importantly, position circadian rhythm integrity and nocturnal light exposure as critically understudied dimensions warranting dedicated mechanistic investigation.

## Introduction

Accelerometer-derived behavioral phenotype provides a multidimensional characterization of human diurnal behavior that extends well beyond physical activity, encompassing daytime movement structure, sleep characteristics, circadian rhythms, step accumulation, and light exposure^1-4^. Yet most wearable-based cancer epidemiology studies have focused narrowly on total activity or a limited complement of sleep characteristics, leaving the broader behavioral architecture and its relationship to solid cancer risk incompletely characterized^5, 6^. This gap is particularly consequential for circadian organization and light exposure, both of which are biologically intertwined with canonical hallmarks of cancer, including DNA repair fidelity, hormonal regulation, and immune surveillance^7-11^. Despite compelling mechanistic plausibility, these domains remain far less developed than movement-volume measures in wearable epidemiology, representing a critical and underexplored frontier.

A five-domain wearable framework is therefore not merely additive but conceptually necessary. Because behavioral variables are inherently intercorrelated, they likely reflect an integrated “behavioral phenotype” that collectively encodes the systemic metabolic and chronobiological milieu in which tumors initiate and progress^11-13^. Evaluating these domains in isolation risks misattributing shared variance and obscuring the true architecture of behavioral cancer risk^12^.

This study was designed as an accelerometer-derived behavioral exposome analysis of incident cancer outcome. Rather than isolating individual metrics, we evaluated the full wearable phenotype as a dynamic, five-domain system^1, 2^. Incident solid cancers were prioritized as the primary aggregate endpoint acknowledging that while cancer comprises a collection of heterogeneous diseases, many solid cancers share common behavioral risk determinants related to energy balance, systemic inflammation, and circadian disruption^5, 7, 8, 10, 14^. This hierarchical design supports an atlas-style interpretation^15, 16^. Identifying a broad aggregate signal first allows detection of systemic behavioral vulnerabilities. Subsequent site-specific analyses then characterize the differential sensitivity of individual organ systems to distinct behavioral domains^1, 16^. Of particular interest is the potential link between diurnal light exposure and hormone-dependent or metabolically sensitive cancers.

Our objectives were threefold. First, to characterize the accelerometer-wide pattern of associations between five-domain behavioral measures and the risk of incident cancers, thereby establishing a baseline behavioral risk profile for cancer. Second, to identify which site-specific cancers most strongly mirror or diverge from these aggregate patterns. Third, and most critically, to determine whether circadian organization and light exposure metrics contribute independent, non-redundant information beyond the dominant movement-volume and fragmentation architecture^12^. This question carries direct implications for both cancer surveillance and our mechanistic understanding of chronobiological carcinogenesis. By moving from a pan-cancer perspective to site-specific resolution, this analysis provides a high-dimensional behavioral map of cancer risk^16^. The central question is whether particular behavioral domains act as universal modifiers of cancer risk or, alternatively, as site-specific oncogenic triggers.

## Methods

### Study population

We utilized the UK Biobank accelerometry sub-cohort^3, 17^. Between 2013 and 2015, a subset of participants were invited to wear a wrist-worn Axivity AX3 (Newcastle upon Tyne, UK) triaxial accelerometer for 7 days^17^. The accelerometer was set at 100Hz with acceleration range ±8 g^3^. We excluded participants with invalid accelerometry data or loss to follow up. For each specific cancer endpoint, prevalent cases at the time of accelerometer assessment were excluded from the at-risk set. For the primary outcome which is solid cancer, a total of 89,080 participants remained after excluding 6,679 individuals with prevalent solid cancers.

### Accelerometer processing and five-domain framework

Accelerometer data were processed using GGIR (v3.3-4) in R^18, 19^. Light data were first extracted at 100 Hz and subsequently calibrated and converted from raw counts to Lux units following previously validated methods^4, 20-22^. Raw 100 Hz CWA files were downsampled to 15 Hz, and the Verisense step algorithm with v2 parameter^23, 24^ was used to derive step count, bout characteristics, and cadence. Incidental steps, purposeful steps, light and MVPA steps were additionally derived according to previously literature^25^.

A valid wear day was defined as comprising at least 16 hours of data, inclusive of the sleep period. Non-wear detection was performed using the GGIR algorithm. The horizontal angle method^26^ was used to identify the sleep period time (SPT), from which sleep characteristics were subsequently derived. Physical activity intensity was classified using established cut-point thresholds: light physical activity (40-100 mg), moderate activity (100-400 mg), and vigorous activity (>400 mg)^27, 28^. MVPA was defined as bouts of at least 5-10 minutes in duration, with a 80% inclusion criterion applied within each bout^29^.

To capture the fine-grained temporal structure of behavior, fragmentation analysis was applied separately at two levels: the waking period (to characterize daytime physical activity patterning) and the full accelerometry window (to capture sleep-wake fragmentation). For each level, a comprehensive suite of fragmentation metrics was derived. Fragment counts^30^ were quantified for inactivity, total physical activity, LIPA, and MVPA bouts, alongside the corresponding fragment rates per minute spent in each state. Transition probabilities^31^ were computed between key behavioral states, including inactivity-to-activity, inactivity-to-LIPA, and inactivity-to-MVPA transitions. Bout duration characteristics were summarized using the mean, standard deviation, coefficient of variation^32^, and Gini index^33^ for both inactivity and activity fragments, providing indices of both central tendency and distributional inequality. Finally, the scaling properties of bout duration distributions were characterized by fitting a power-law model to inactivity and activity fragment durations, yielding the power-law exponent (alpha) and associated parameters (x0.5 and W0.5),^30, 33^ which index the shape and tail behavior of the distribution.

Circadian rhythm metrics were derived using both parametric and non-parametric approaches, applied across the full accelerometry window. Cosinor and extended cosinor models^34^ were fitted to the acceleration time series to derive the Mesor (midline-estimating statistic of rhythm), amplitude, and acrophase. The most active 10-hour window (M10) and least active 5-hour window (L5) were identified, and Relative Amplitude (RA) was computed to index the overall strength and regularity of the rest-activity rhythm. Interdaily Stability (IS) and Intradaily Variability (IV)^31, 35, 36^ were calculated to quantify the alignment of behavioral patterns with the 24-hour solar cycle and the degree of hourly fragmentation within the rest-activity sequence, respectively. The Self-Similarity Parameter (SSP)^37^ and Activity Balance Index (ABI)^31^ were derived via Detrended Fluctuation Analysis (DFA) to characterize the long-range correlation structure and intrinsic complexity of the acceleration signal. Finally, the first-order autocorrelation of multi-day activity patterns (Phi)^31, 38^ was estimated using a first-order autoregressive model to capture the day-to-day temporal consistency of the circadian behavioral system.

### Cancer outcome definition

Cancer outcomes were ascertained from incident cancer registry diagnoses using ICD-10 codes, categorized according to previous literature^39^. The primary aggregate outcome was incident solid cancer, defined as any first primary solid cancer (ICD-10 codes C00-C80, excluding non-melanoma skin cancer C44). Secondary outcomes comprised 16 prespecified site-specific solid cancers: prostate (C61), breast (C50), colorectal (C18, C19, and C20), lung (C34), melanoma (C43), uterine (C54, C55), kidney (C64, C65), bladder (C67), pancreatic (C25), ovarian (C56), esophageal (C15), brain (C71), stomach (C16), liver (C22), soft tissue (C46, C47, C48, C49), and thyroid (C73) cancers. Hematological malignancies including lymphoma (C81, C82, C83, C84, C85, C86, C88), leukemia (C91, C92, C93, C94, C95), and multiple myeloma (C90.0) were additionally examined as exploratory outcomes. For each outcome, baseline prevalence was determined to enable exclusion of pre-existing diagnoses, and participant-level event indicators, dates of first incident diagnosis, and total person-time at risk were derived for survival analysis.

### Statistical analysis

We used Cox proportional hazards models with age as timescale to evaluate the associations between accelerometer-derived behavioral exposomes and incident cancer outcomes. The primary analytic tier (Minimal Model) adjusted for sex (male/female), ethnicity (White/other), Townsend Deprivation Index (continuous), smoking status (never/past/current), alcohol consumption (never/past/current), family history of cancer (any first-degree history of prostate, breast, lung, or colorectal cancer versus none), and seasonality of accelerometer assessment (winter, spring, summer, or autumn). To assess the robustness of the observed associations, four subsequent sensitivity tiers were evaluated. These tiers sequentially incorporated additional adjustments for: (1) adiposity and metabolic factors (diabetes and BMI); (2) mean acceleration (in mg, to isolate exposure-specific associations independent of overall activity volume); (3) shift-work history (yes/no); and (4) accelerometry measurement quality (wear-time percentage and number of valid days).

For each exposure-outcome pair, effect estimates were expressed as hazard ratios (HR) per one standard deviation increment in the continuous exposure, complemented by tertile-based comparisons and tests for linear trend across tertile medians. The threshold for nominal statistical significance was set at two-sided P < 0.05. Comprehensive results for each endpoint are reported in Supplementary Tables S1-S20, with a pan-cancer synthesis and a summary of recurrent nominal signals provided in Supplementary Tables S21 and S22, respectively. All analyses were conducted in R version 4.5.3^40^.

## Results

### Study population

The final analytic cohort comprised 89,080 participants at risk for incident cancer, following the exclusion of 6,679 individuals with prevalent cancer diagnoses at baseline. Over a median follow-up of 8.39 years, 6,598 incident solid cancer events were recorded. Among site-specific outcomes, event counts were highest for prostate (n = 1,792), breast (n = 1,272), and colorectal (n = 775) cancers. Substantial numbers of incident events were also observed for melanoma (n = 439), lung cancer (n = 413), and lymphoma (n = 345). Additional evaluable endpoints included uterine (n = 202), kidney (n = 198), pancreatic (n = 196), leukemia (n = 184), bladder (n = 156), esophageal (n = 145), ovarian (n = 141), and multiple myeloma (n = 133) cancers.

### Pan-Cancer AWAS Atlas: Primary Outcomes

In the Minimal Model, incident solid cancer as an aggregate endpoint demonstrated a broad signal burden across the behavioral exposome. The strongest inverse associations were observed for higher overall acceleration (HR per SD: 0.91, 95% CI: 0.89-0.94; P = 7.0 × 10^−11^), greater daily step count (HR: 0.93, 95% CI: 0.90-0.95; P = 3.2 × 10^−9^), and longer moderate-intensity activity duration (HR: 0.92, 95% CI: 0.90-0.94; P = 7.7 × 10^−10^). Conversely, inactivity fragmentation metrics were associated with elevated risk; the x0.5 parameter of inactivity fragment duration distributions, indexing the characteristic timescale of sedentary bouts, was among the most prominent positive associations (HR: 1.08, 95% CI: 1.05-1.10; P = 5.1 × 10^−11^). Although the aggregate signal architecture was dominated by movement volume and fragmentation, circadian metrics featured prominently as well, with cosinor mesor, relative amplitude, and related rhythm-strength indices showing recurrent inverse associations with solid cancer risk.

Tertile analyses supported dose-response gradients across key exposures. Participants in the highest versus lowest tertile of overall acceleration had markedly lower all-solid-cancer risk (HR: 0.84, 95% CI: 0.79-0.89; P-trend = 3.6 × 10^−8^), with analogous gradients observed for total moderate-intensity activity duration and daily step count. Conversely, participants in the highest tertiles of selected inactivity fragmentation metrics exhibited elevated risk relative to the lowest tertiles. Adjustment for adiposity and metabolic factors attenuated several associations, but the principal movement-volume signal remained materially unchanged. In the Independent Pattern Model—which additionally adjusted for mean acceleration—selected structured activity and fragmentation signals retained nominal statistical significance, indicating that a portion of the primary signal was not fully explained by overall activity volume alone.

### Secondary outcomes

Site-specific analyses revealed heterogeneous but interpretable patterns across tumor sites. Lung cancer exhibited the strongest and most coherent inverse activity profile, with reduced risk associated with higher moderate-intensity activity duration, greater overall acceleration, and higher daily step count. Breast cancer demonstrated reproducible inverse associations with structured moderate-to-vigorous physical activity metrics, while colorectal cancer showed a narrower but directionally concordant pattern centered on structured MVPA accumulation. Among lower-count endpoints, two findings were particularly noteworthy: a selective association between nocturnal light exposure and pancreatic cancer risk, and pronounced inactivity-related associations for uterine cancer in the Minimal Model that were substantially attenuated following adjustment for adiposity and metabolic factors.

Across site-specific outcomes, recurrent nominal associations were dominated by physical activity volume and fragmentation metrics; however, the circadian and light exposure domains contributed important, non-redundant information. The most consistently recurring signals across outcomes included mean inactivity fragment duration, x0.5 inactivity fragment parameters, relative amplitude, cosinor amplitude, and selected inactivity-to-MVPA transition probabilities. The recurrence of rhythm-strength metrics across multiple tumor sites supports circadian organization as a substantive and independent component of the behavioral cancer risk atlas, rather than a peripheral or epiphenomenal domain. Light exposure did not emerge as a major driver of aggregate solid cancer risk, but demonstrated a more concentrated and potentially site-specific contribution— most prominently for pancreatic cancer—underscoring the relevance of the light domain for site-specific etiological prioritization. Comprehensive outcome-specific results are reported in Supplementary Tables S1-S20, the cross-cancer signal summary in Supplementary Table S21, and the recurrent-signal synthesis in Supplementary Table S22.

## Discussion

### Principal findings

Across this five-domain accelerometer-wide association study (AWAS)^6, 41, 42^, the primary all-solid-cancer endpoint showed a consistent phenotype: higher movement volume and greater moderate-intensity activity were associated with lower incidence, whereas greater inactivity accumulation and several inactivity-fragmentation metrics were associated with higher incidence. This broad pattern persisted directionally across model tiers, with partial attenuation after adiposity/metabolic adjustment and selective persistence for structured activity metrics in the Independent Pattern model.

### Interpretation

These findings support the interpretation of accelerometer profiles as reflecting an integrated behavioral exposome rather than collections of isolated single-variable effects^1, 13, 43^. Movement volume, movement structure, and inactivity burden appear to capture overlapping yet complementary dimensions of cancer-relevant behavior. The broad all-solid-cancer signal architecture was dominated by physical activity and fragmentation measures, while light and sleep variables contributed more selectively to particular endpoints. The site-specific heterogeneity observed, most notably for lung, breast, colorectal, pancreatic, and uterine cancers, argues for a hierarchical behavioral exposome interpretation in which shared movement phenotypes coexist with outcome-specific behavioral risk profiles.

The circadian findings warrant explicit emphasis, as they extend the interpretive framework beyond simple activity dose. Metrics such as cosinor mesor, relative amplitude, and related rhythm-strength indices capture the organization, timing, and day-night contrast of the behavioral cycle, rather than the quantity of movement accumulated alone^5, 9, 44^. Their emergence among the leading and most recurrent associations suggests that cancer-relevant behavioral exposure may depend not only on how much movement is accrued, but on how behavior is distributed and structured across the 24-hour day^33^. This is among the more distinctive contributions of the present analysis, and provides empirical support for evaluating the full five-domain behavioral exposome rather than movement volume in isolation^1^.

The pancreatic cancer finding warrants specific attention, as it diverges meaningfully from the dominant pan-cancer pattern. Whereas most solid cancers were primarily associated with movement volume and fragmentation metrics, pancreatic cancer demonstrated a localized, robust association with higher nocturnal light exposure that persisted across sensitivity models. This selective vulnerability represents one of the most novel and conceptually distinctive observations to emerge from the AWAS framework^6, 16, 41, 42^.

The biological plausibility for a specific light-pancreas axis is compelling and multifaceted. The pancreas is an exquisitely rhythm-dependent organ^45^. Diurnal light exposure is the primary environmental suppressor of melatonin, a hormone critical for maintaining glucose homeostasis via receptors on pancreatic islets^45^. Light-induced melatonin suppression^46, 47^, often coupled with altered feeding timing such as late-night eating, can induce profound circadian misalignment between the central pacemaker and pancreatic peripheral clocks^4^. Chronic temporal perturbation of this kind exacerbates hyperinsulinemia, insulin resistance, and local oxidative stress^48^, a well-established metabolic and inflammatory precursor to pancreatic carcinogenesis^45, 49^.

Furthermore, this distinction underscores the value of a multidimensional exposome approach^6, 16, 41, 42^. It suggests that while physical activity acts as a diffuse, systemic modifier of cancer risk, chronobiological exposures such as nocturnal light may operate as site-specific modulators of cancer risk in metabolically sensitive organs^14^. However, given the modest event count for incident pancreatic cancer (n = 196) in this cohort, we interpret this site-specific signal with appropriate caution. Rather than drawing definitive causal conclusions, we propose this nocturnal light-pancreatic cancer association as a high-priority, mechanistically grounded hypothesis for targeted replication in independent cohorts and functional chronobiology studies.

### Strengths and limitations

Key strengths of this study include objective, device-based exposure assessment, simultaneous evaluation of five behavioral domains within a unified analytic framework, and a tiered modeling strategy that systematically distinguishes minimally adjusted associations from those additionally accounting for adiposity, metabolic factors, and independent activity patterns. The hierarchical endpoint structure, with all incident solid cancers as the primary outcome and prespecified site-specific cancers as secondary outcomes, effectively separates broad, high-power patterns from lower-power exploratory findings.

Important limitations nonetheless remain. Associations were evaluated at nominal significance thresholds across a high-dimensional and internally correlated exposure set; accordingly, multiplicity and shared variance across exposures may inflate the apparent signal burden. Accelerometer phenotypes were derived from a single baseline measurement window and may not reflect long-term behavioral patterns. Event counts varied substantially across site-specific outcomes, and findings for lower-count endpoints should be interpreted with commensurate caution. As with any observational study design, residual confounding and the possibility of reverse causation cannot be excluded. Site-specific findings particularly those for less common cancers are best regarded as hypothesis-generating and in need of independent replication.

**Figure 1.**
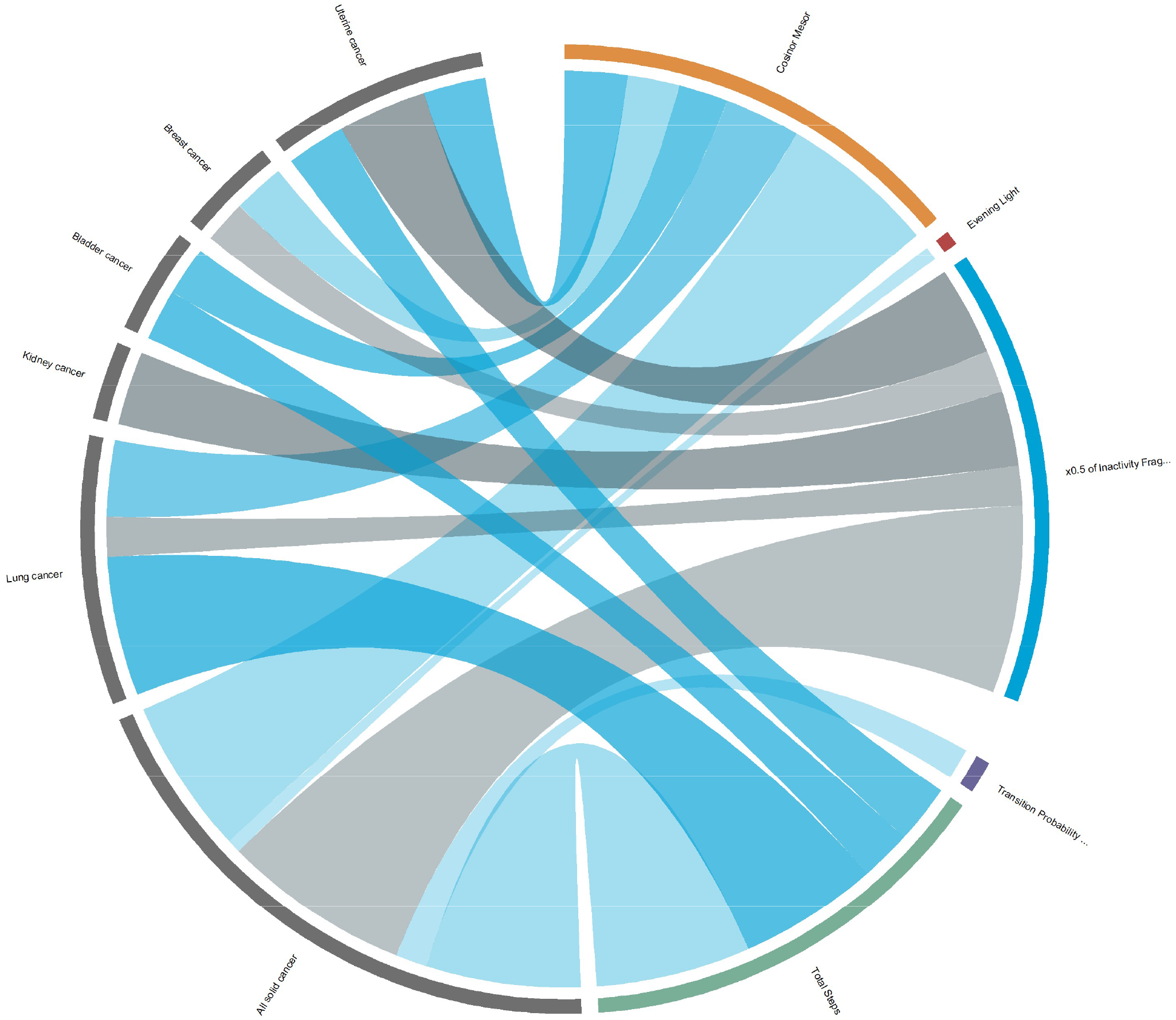
Mapping the behavioral exposome to incident cancer risk. Chord diagram illustrating the associations between the five-domain accelerometer-derived behavioral exposome and incident cancer risk. Selected leading behavioral metrics associated with the primary aggregate endpoint (all solid cancer) are mapped against their specific associations with recurrent site-specific cancers. Exposure variables are categorized according to the five-domain framework (e.g., circadian rhythms, light exposure, inactivity fragmentation, and movement volume), with the width of the connecting ribbons weighted by the relative signal strength of each association.

**Figure 2.**
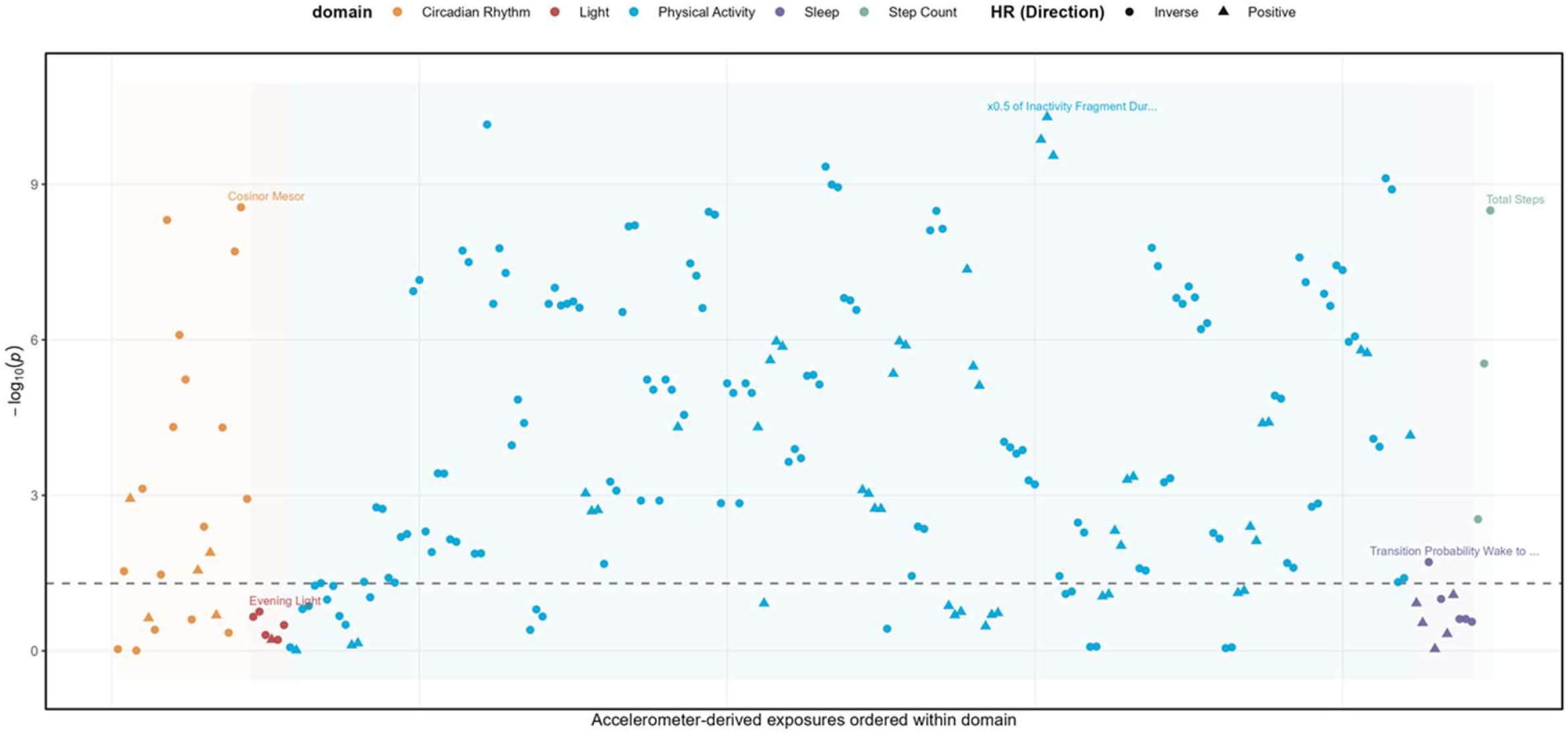
Accelerometer-wide association study (AWAS) of incident solid cancer. Accelerometer-wide association study (AWAS) atlas of incident solid cancer risk. The plot visualizes the baseline associations (Minimal Model) between the five-domain behavioral exposome and the primary aggregate endpoint (all solid cancers). Individual accelerometer-derived metrics are grouped along the x-axis according to their respective behavioral domains (Circadian Rhythm, Light, Physical Activity, Sleep, and Step Count). The y-axis represents the -log10(p) value for each association, with the horizontal dashed line indicating the nominal significance threshold. Point shapes denote the direction of the hazard ratio (HR), distinguishing between inverse (circles, indicating reduced risk) and positive (triangles, indicating elevated risk) associations.

**Figure 3.**
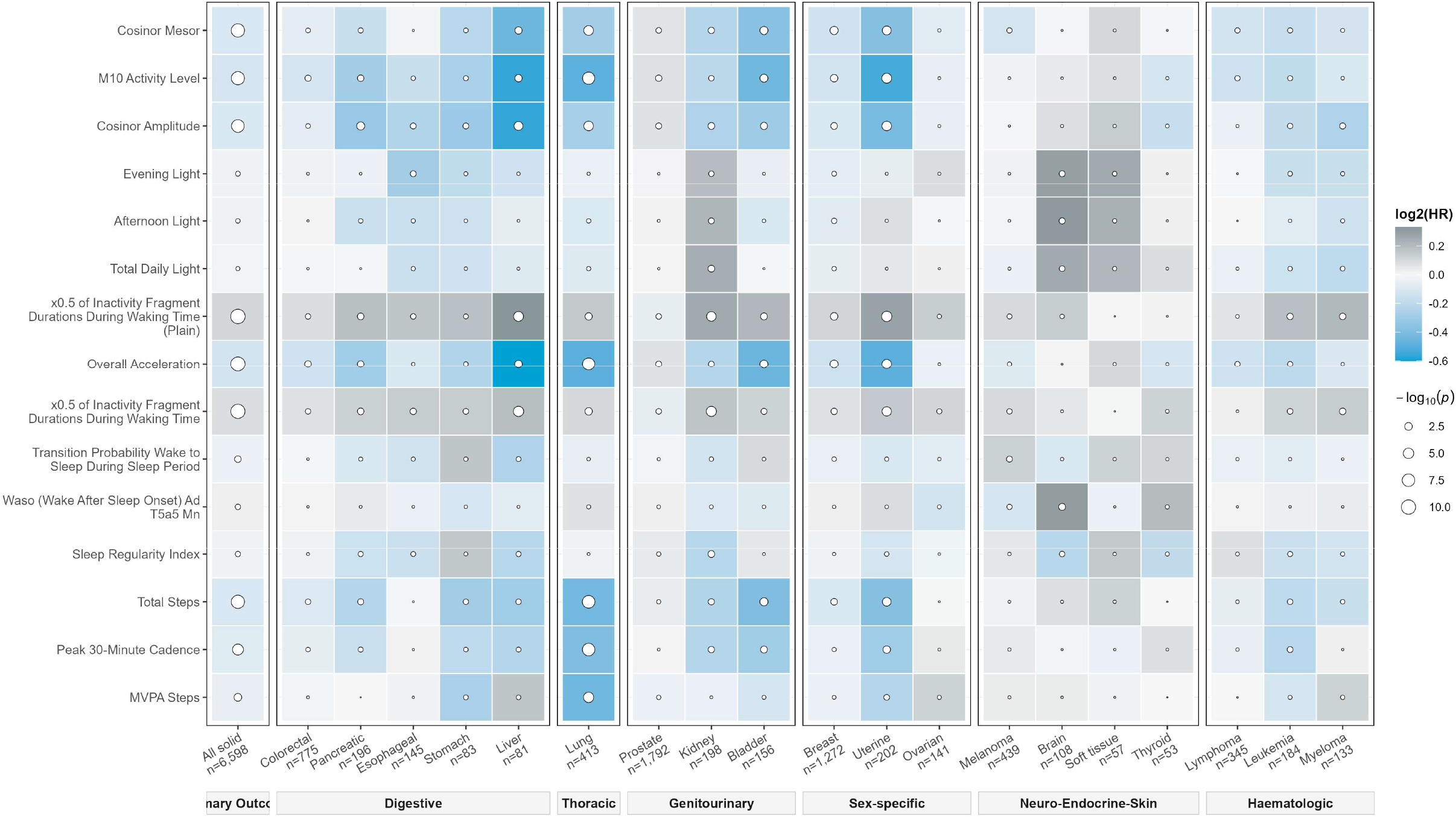
Site-specific association profiles of lead behavioral exposures. Comprehensive AWAS heatmap detailing the risk architecture of the behavioral exposome across incident cancer outcomes. The plot maps recurrent lead exposures against the primary aggregate endpoint (all solid cancers) and secondary site-specific malignancies with at least 51 incident events. Columns are hierarchically grouped by anatomical and systemic categories. Rows display key accelerometer-derived metrics spanning the five behavioral domains. The magnitude and direction of the associations are encoded by tile background color [log2(HR)], while superimposed circle size indicates statistical significance [-log10(p)].

**Figure 4.**
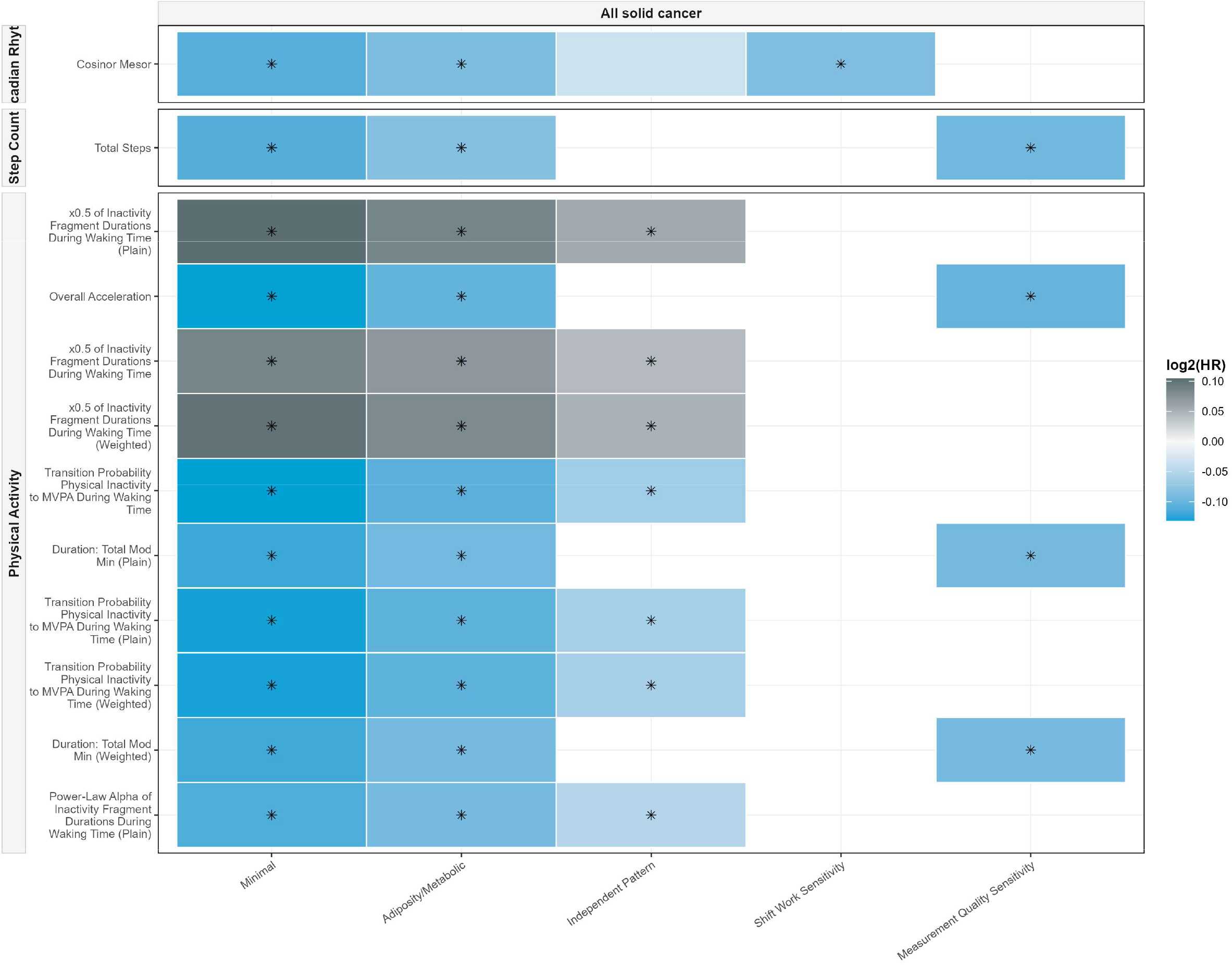
Robustness of lead behavioral signals across tiered sensitivity models. Robustness of the behavioral exposome associations with incident solid cancer across tiered sensitivity models. The tile plot illustrates the stability of lead accelerometer-derived signals for the primary aggregate endpoint (all solid cancers) across five sequentially adjusted analytical tiers: Minimal, Adiposity/Metabolic, Independent Pattern, Shift Work Sensitivity, and Measurement Quality Sensitivity models. Exposures are vertically faceted by their respective behavioral domains (e.g., Circadian Rhythm, Step Count, and Physical Activity). Tile background color encodes the magnitude and direction of the hazard ratio [log2(HR)],, while superimposed asterisks (*) denote nominal statistical significance (P<0.05).

**Table 1.** Cohort Characteristics and Outcome Counts.

| Characteristic | Overall | Non-cancer | Incident Cancer |
| --- | --- | --- | --- |
| N | 89080 | 82482 | 6598 |
| Age at accelerometry, years | 62.1 (7.9) | 61.9 (7.9) | 65.3 (6.8) |
| Male, n (%) | 39133 (43.9) | 35579 (43.1) | 3554 (53.9) |
| White, n (%) | 85974 (96.5) | 79525 (96.4) | 6449 (97.7) |
| Townsend deprivation index | -1.7 (2.8) | -1.7 (2.8) | -1.8 (2.8) |
| Body mass index, kg/m<sup>2</sup> | 26.7 (4.5) | 26.7 (4.5) | 27.2 (4.5) |
| Smoking status, n (%) |  |  |  |
| Never | 51225 (57.5) | 47915 (58.1) | 3310 (50.2) |
| Past | 31668 (35.6) | 28937 (35.1) | 2731 (41.4) |
| Current | 6187 (6.9) | 5630 (6.8) | 557 (8.4) |
| Alcohol status, n (%) |  |  |  |
| Never | 2576 (2.9) | 2398 (2.9) | 178 (2.7) |
| Past | 2439 (2.7) | 2255 (2.7) | 184 (2.8) |
| Current | 84065 (94.4) | 77829 (94.4) | 6236 (94.5) |
| Diabetes, n (%) | 3059 (3.4) | 2782 (3.4) | 277 (4.2) |
| Family history of cancer, n (%) | 33402 (37.5) | 30637 (37.1) | 2765 (41.9) |
| Accelerometry season, n (%) |  |  |  |
| Spring | 20386 (22.9) | 18853 (22.9) | 1533 (23.2) |
| Summer | 23427 (26.3) | 21716 (26.3) | 1711 (25.9) |
| Autumn | 26472 (29.7) | 24539 (29.8) | 1933 (29.3) |
| Winter | 18795 (21.1) | 17374 (21.1) | 1421 (21.5) |
| Incident cancer outcomes, n |  |  |  |
| All malignant solid neoplasms | 6598 |  |  |
| Prostate cancer | 1792 |  |  |
| Breast cancer | 1272 |  |  |
| Colorectal cancer | 775 |  |  |
| Melanoma | 439 |  |  |
| Lung cancer | 413 |  |  |
| Lymphoma | 345 |  |  |
| Uterine cancer | 202 |  |  |
| Kidney cancer | 198 |  |  |
| Pancreatic cancer | 196 |  |  |
| Leukemia | 184 |  |  |
| Bladder cancer | 156 |  |  |
| Esophageal cancer | 145 |  |  |
| Ovarian cancer | 141 |  |  |
| Multiple myeloma | 133 |  |  |
| Brain cancer | 108 |  |  |
| Stomach cancer | 83 |  |  |
| Liver cancer | 81 |  |  |
| Soft tissue cancer | 57 |  |  |
| Thyroid cancer | 53 |  |  |

**Table 2.** Association between accelerometer-derived Behavioral exposome and incident all solid cancer risk.

| Domain | Exposure | Cases T1 | Cases T2 | Cases T3 | HR T1 | HR T2 | HR T3 | HR per SD | P trend |
| --- | --- | --- | --- | --- | --- | --- | --- | --- | --- |
| Physical Activity | x0.5 of Inactivity Fragment Durations During Waking Time (Plain) | 1903 | 2155 | 2540 | Reference | 1.03 [0.97;1.10] | 1.15 [1.08;1.22] | 1.08 [1.05;1.10] | 2.25e-6 |
| Physical Activity | Overall Acceleration | 2568 | 2185 | 1845 | Reference | 0.92 [0.87;0.98] | 0.84 [0.79;0.89] | 0.91 [0.89;0.94] | 3.62e-8 |
| Physical Activity | x0.5 of Inactivity Fragment Durations During Waking Time | 1893 | 2159 | 2546 | Reference | 1.04 [0.98;1.10] | 1.15 [1.08;1.22] | 1.06 [1.04;1.08] | 3.34e-6 |
| Physical Activity | x0.5 of Inactivity Fragment Durations During Waking Time (Weighted) | 1906 | 2154 | 2538 | Reference | 1.03 [0.97;1.09] | 1.14 [1.08;1.22] | 1.07 [1.05;1.10] | 3.42e-6 |
| Physical Activity | Transition Probability Physical Inactivity to MVPA During Waking Time | 2659 | 2148 | 1791 | Reference | 0.89 [0.84;0.94] | 0.85 [0.80;0.91] | 0.91 [0.89;0.94] | 1.03e-6 |
| Physical Activity | Total Moderate Physical Activity Duration (Plain) | 2541 | 2158 | 1899 | Reference | 0.92 [0.87;0.97] | 0.86 [0.81;0.91] | 0.92 [0.90;0.94] | 1.2e-6 |
| Physical Activity | Transition Probability Physical Inactivity to MVPA During Waking Time (Plain) | 2655 | 2158 | 1785 | Reference | 0.90 [0.85;0.95] | 0.85 [0.80;0.91] | 0.91 [0.89;0.94] | 7.33e-7 |
| Physical Activity | Transition Probability Physical Inactivity to MVPA During Waking Time (Weighted) | 2648 | 2160 | 1790 | Reference | 0.90 [0.85;0.95] | 0.85 [0.80;0.91] | 0.91 [0.89;0.94] | 1.11e-6 |
| Physical Activity | Total Moderate Physical Activity Duration (Weighted) | 2541 | 2158 | 1899 | Reference | 0.92 [0.87;0.98] | 0.86 [0.81;0.91] | 0.92 [0.90;0.95] | 1.16e-6 |
| Circadian Rhythm | Cosinor Mesor | 2533 | 2160 | 1905 | Reference | 0.92 [0.86;0.97] | 0.87 [0.82;0.92] | 0.93 [0.90;0.95] | 4.77e-6 |
| Step Count | Total Steps | 2493 | 2142 | 1963 | Reference | 0.92 [0.86;0.97] | 0.87 [0.82;0.93] | 0.93 [0.90;0.95] | 6.46e-6 |
| Physical Activity | Power-Law Alpha of Inactivity Fragment Durations During Waking Time (Plain) | 2566 | 2153 | 1879 | Reference | 0.89 [0.84;0.94] | 0.86 [0.81;0.91] | 0.92 [0.90;0.95] | 4.25e-7 |

**Table 3.**
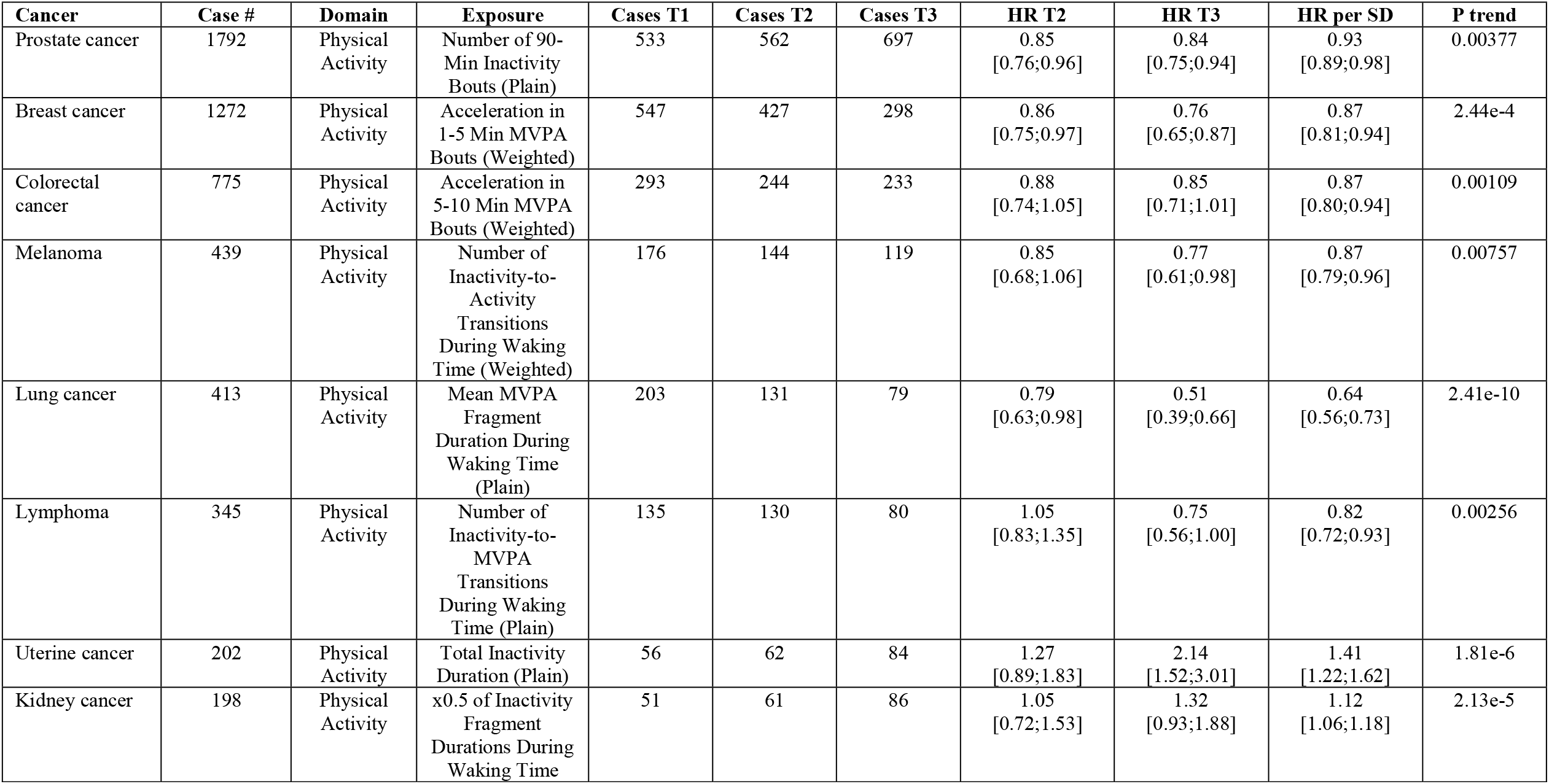
Association between accelerometer-derived Behavioral exposome and incident site-specific solid cancers and hematological malignancies risk.

## Supporting information

Table S1: All solid cancer, incident n=6,598

able S2: Prostate cancer, incident n=1,792

Table S3: Breast cancer, incident n=1,272

Table S4: Colorectal cancer, incident n=775

Table S5: Melanoma, incident n=439

Table S6: Lung cancer, incident n=413

Table S7: Lymphoma, incident n=345

Table S8: Uterine cancer, incident n=202

Table S9: Kidney cancer, incident n=198

Table S10: Pancreatic cancer, incident n=196

Table S11: Leukemia, incident n=184

Table S12: Bladder cancer, incident n=156

Table S13: Esophageal cancer, incident n=145

Table S14: Ovarian cancer, incident n=141

Table S15: Multiple myeloma, incident n=133

Table S16: Brain cancer, incident n=108

Table S17: Stomach cancer, incident n=83

Table S18: Liver cancer, incident n=81

Table S19: Soft tissue cancer, incident n=57

Table S20: Thyroid cancer, incident n=53

Cross-cancer AWAS summary of completed outcomes

Shared nominally significant behavioral signals across site-specific malignancies

## Data Availability

UK Biobank data are available at https://www.ukbiobank.ac.uk/.

## Ethical approval

The North West Multi-Centre Research Ethics Committee approved the protocol of UK Biobank (REC reference: 11/NW/0382), and all participants provided informed consent before data collection

## Data Sharing

UK Biobank data are available at https://www.ukbiobank.ac.uk/.

## Author Contributions

J.A.B. designed the study, performed statistical analyses.

M.N.C.C. interpreted results and wrote the manuscript.

## Figures and tables

**Extended Data Figure 1.**
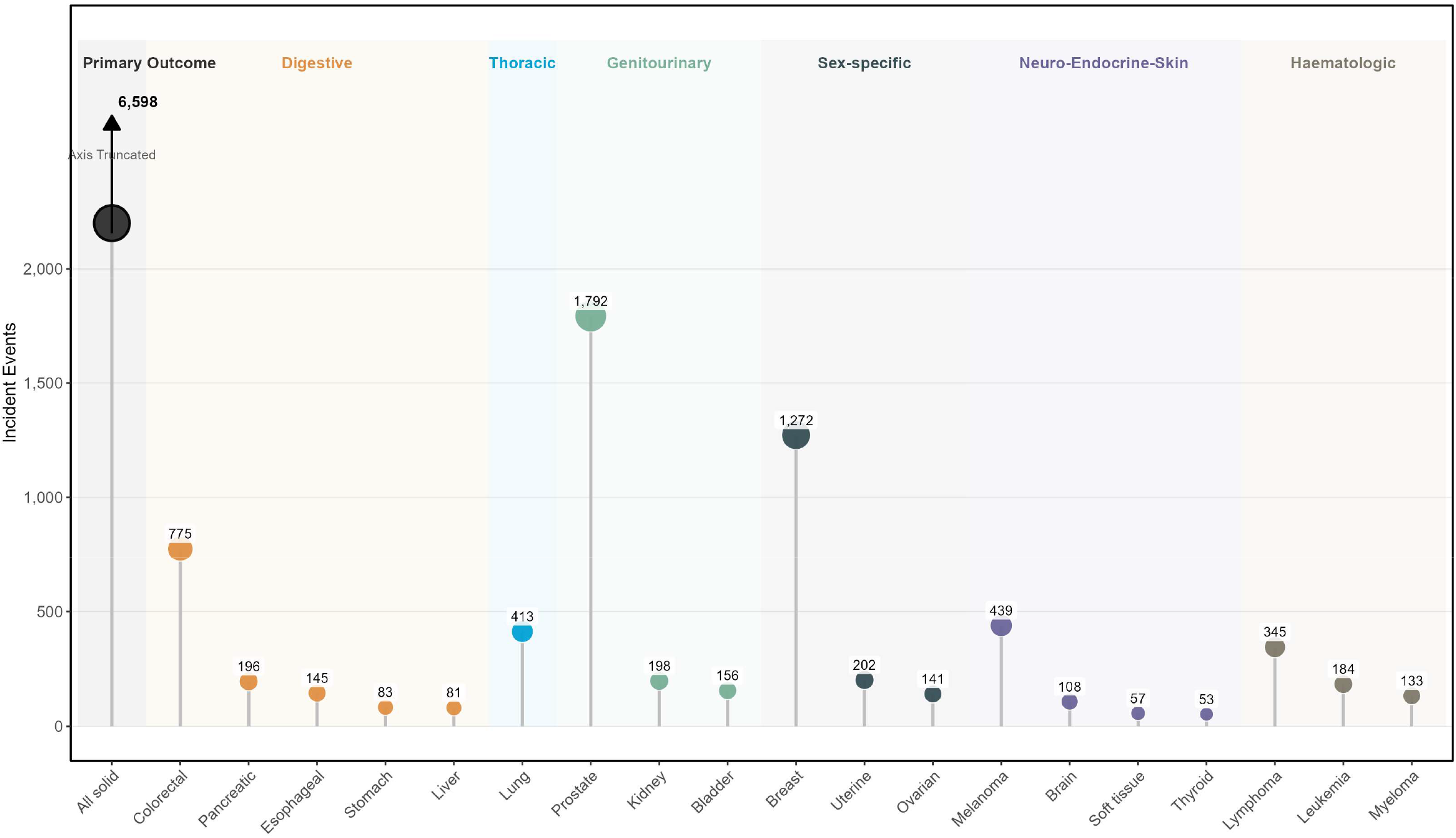
Hierarchical Distribution of Incident Cancer Events. Baseline incidence architecture across prespecified cancer outcomes. The lollipop plot illustrates the distribution of incident events for malignancies with at least 50 cases within the analytic cohort. Site-specific outcomes (x-axis) are hierarchically organized by anatomical and systemic categories. To preserve the visual resolution of lower-count malignancies, the primary aggregate endpoint (all solid cancers, n = 6,598) is displayed using a truncated y-axis.

**Extended Data Figure 2.**
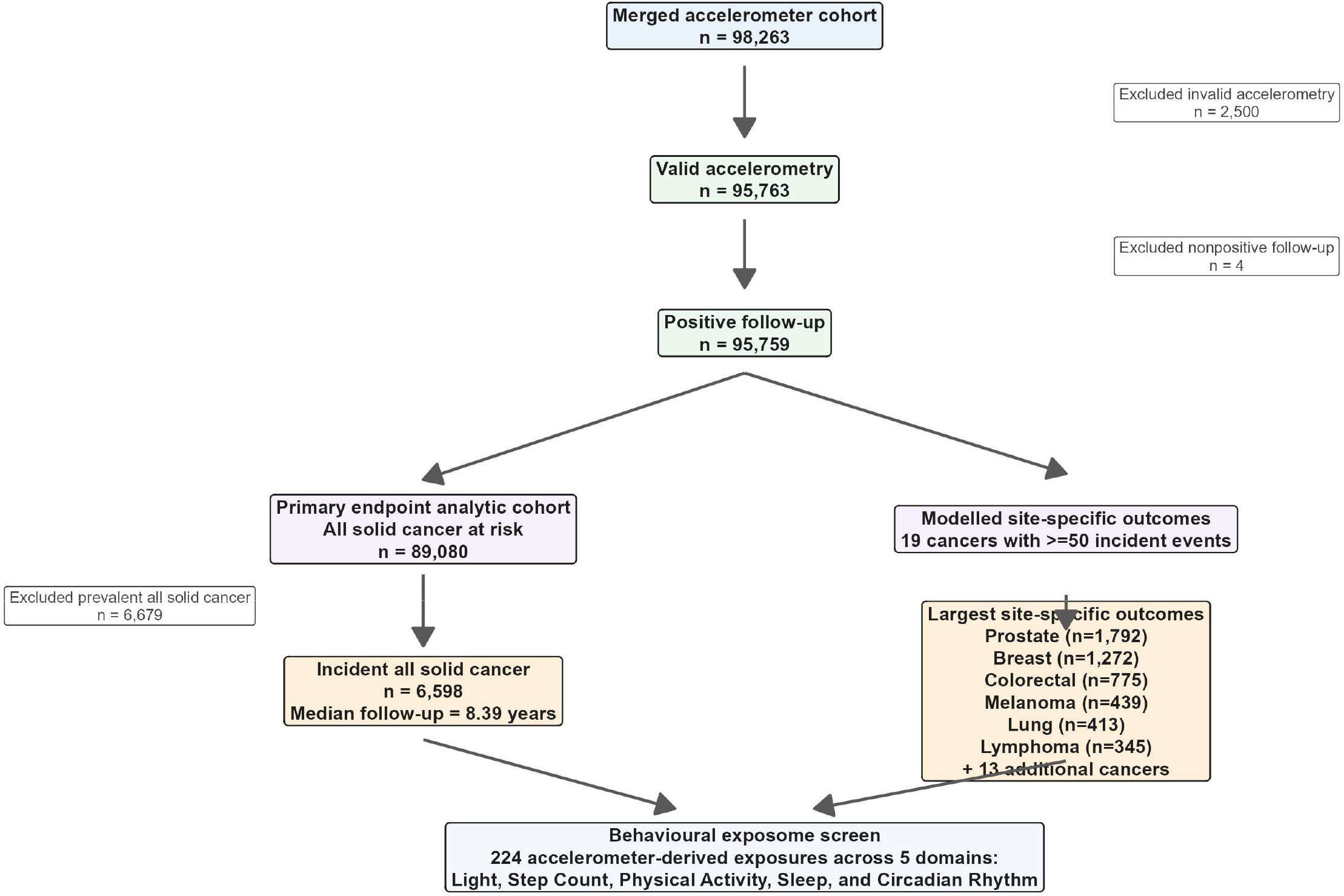
Study Flow Chart.

**Extended Data Figure 3.**
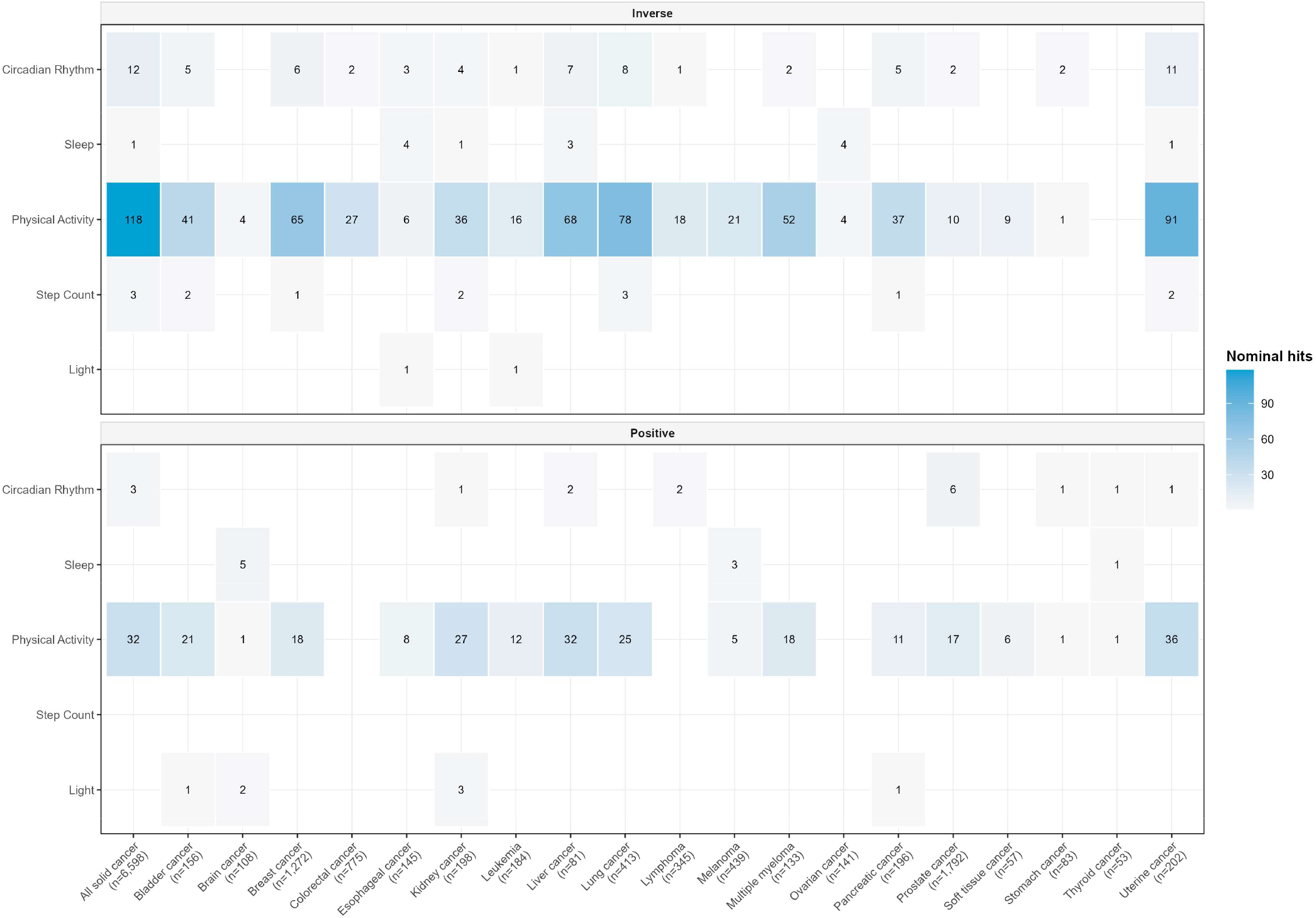
AWAS Signal Burden Architecture Across the Behavioral Exposome. Heatmap illustrating the cumulative nominal signal burden across the five-domain behavioral exposome. The matrix quantifies the total number of nominally significant associations (P < 0.05) identified between specific accelerometer-derived metrics and modeled cancer outcomes. The plot is stratified by the direction of the hazard ratio: inverse associations indicating reduced risk (top panel) and positive associations indicating elevated risk (bottom panel). Both the numeric values and the blue color intensity within each tile denote the absolute count of significant “hits” for a given behavioral domain and cancer pair.

## Supplementary Tables

The full completed paper-2 model outputs are provided below.

- Table S1: All solid cancer, incident n=6,598 (table_s01_all_solid_cancer_full_results.csv)
- Table S2: Prostate cancer, incident n=1,792 (table_s02_prostate_full_results.csv)
- Table S3: Breast cancer, incident n=1,272 (table_s03_breast_full_results.csv)
- Table S4: Colorectal cancer, incident n=775 (table_s04_colorectal_full_results.csv)
- Table S5: Melanoma, incident n=439 (table_s05_melanoma_full_results.csv)
- Table S6: Lung cancer, incident n=413 (table_s06_lung_full_results.csv)
- Table S7: Lymphoma, incident n=345 (table_s07_lymphoma_full_results.csv)
- Table S8: Uterine cancer, incident n=202 (table_s08_uterine_full_results.csv)
- Table S9: Kidney cancer, incident n=198 (table_s09_kidney_full_results.csv)
- Table S10: Pancreatic cancer, incident n=196 (table_s10_pancreatic_full_results.csv)
- Table S11: Leukemia, incident n=184 (table_s11_leukemia_full_results.csv)
- Table S12: Bladder cancer, incident n=156 (table_s12_bladder_full_results.csv)
- Table S13: Esophageal cancer, incident n=145 (table_s13_esophageal_full_results.csv)
- Table S14: Ovarian cancer, incident n=141 (table_s14_ovarian_full_results.csv)
- Table S15: Multiple myeloma, incident n=133 (table_s15_multiplemyeloma_full_results.csv)
- Table S16: Brain cancer, incident n=108 (table_s16_brain_full_results.csv)
- Table S17: Stomach cancer, incident n=83 (table_s17_stomach_full_results.csv)
- Table S18: Liver cancer, incident n=81 (table_s18_liver_full_results.csv)
- Table S19: Soft tissue cancer, incident n=57 (table_s19_softtissue_full_results.csv)
- Table S20: Thyroid cancer, incident n=53 (table_s20_thyroid_full_results.csv)
- Table S21: Cross-cancer AWAS summary of completed outcomes (table_s21_cross_cancer_AWAS_summary.csv)
- Table S22: Shared nominally significant behavioral signals across site-specific malignancies (table_s22_shared_signal_summary.csv)

